# Efficacy and longevity of immune response to 3^rd^ COVID-19 vaccine and effectiveness of a 4^th^ dose in severely immunocompromised patients with cancer

**DOI:** 10.1101/2022.07.05.22277281

**Authors:** Astha Thakkar, Kith Pradhan, Benjamin Duva, Juan Manuel Carreño, Srabani Sahu, Victor Thiruthuvanathan, Sean Campbell, Sonia Gallego, Tushar D Bhagat, Johanna Rivera, Gaurav Choudhary, Raul Olea, Maite Sabalza, Lauren C. Shapiro, Matthew Lee, Ryann Quinn, Ioannis Mantzaris, Edward Chu, Britta Will, Liise-anne Pirofski, Florian Krammer, Amit Verma, Balazs Halmos

## Abstract

Cancer patients show increased morbidity with COVID-19 and need effective immunization strategies. We demonstrate that a 3^rd^ dose of COVID-19 vaccine leads to seroconversion in 57% of patients that were seronegative after primary vaccination. The immune response is durable as assessed by anti-S antibody titers, T-cell activity and neutralization activity against wild-type SARS-CoV2 and BA1.1.529 at 6 months of follow up. A subset of severely immunocompromised hematologic malignancy patients were unable to mount adequate immune response after the 3^rd^ dose and were treated with a 4^th^ dose in a prospective clinical trial which led to adequate immune-boost in 67% of patients. Low baseline IgM levels and CD19 counts were associated with inadequate seroconversion. Booster doses induced limited neutralization activity against the Omicron variant. These results indicate that vaccine booster-induced immunity is durable in cancer patients and additional doses can further stimulate immunity in a subset of hematologic malignancy patients.

**Statement of significance:** We demonstrate that a 3^rd^ dose of vaccine leads to seroconversion in 57% of negative patients with durable immune responses at 6 months. A 4^th^ dose of vaccine can seroconvert hematologic malignancy patients with higher baseline IgM and CD19 levels.

## Introduction

It is now well-established that coronavirus disease 2019 (COVID-19) in patients with cancer carries a higher morbidity and mortality, especially in patients with hematologic malignancies[1-5]. While overall case fatality has decreased over time, mostly related to the impact of broad vaccinations and improved supportive/antimicrobial management, a higher case fatality rate was noted amongst cancer patients even during the Omicron (B.1.1.529) wave [6-8]. Advanced age, co-morbidities and performance status have emerged as key factors adversely impacting outcomes amongst patients with a cancer diagnosis[9]. Effective vaccines have been developed and authorized by the FDA to combat this pandemic[10, 11]. However, emerging data suggests that despite these vaccines inducing high levels of immunity in the general population, patients with hematologic malignancies have lower rates of seroconversion as defined by severe acute respiratory syndrome coronavirus 2 (SARS-CoV-2) spike antibody (anti-S antibody) titers [12, 13]. Evidence has also suggested that specific therapies, such as anti-CD20 antibodies, BTK-inhibitors and stem cell transplantation (SCT) have an association with lower rates of seroconversion [14-16].

We previously published preliminary results of a study defining notable impacts of a 3^rd^ booster dose of vaccine, demonstrating a more than 50% seroconversion rate amongst patients remaining seronegative after primary vaccinations[17]. Since then, we have completed our entire primary cohort to assess initial responses with a broad array of immunological assays along with now additional significant follow up allowing assessment of key aspects of waning immunity. Importantly, we additionally conducted a trial assessing the efficacy of fourth dose vaccinations amongst a highly immune suppressed group of patients with no or limited response to a prior booster vaccine dose. Here we present results of both key cohorts including results of serological, T cell and neutralization assays as well as correlations with other baseline clinical, treatment and laboratory parameters.

## Methods

### Patient recruitment and follow-up (ClinicalTrials.gov identifier NCT05016622)

#### 3^rd^ dose study

We recruited patients via an informed consent process. Patients were required to be >18 years of age and have a cancer diagnosis either on active treatment or requiring active surveillance. Patients were also required to have received 2 doses of the mRNA COVID-19 vaccine or 1 dose of the adenoviral vaccine prior to enrollment. After drawing baseline labs that included spike antibody, a sample for T-cell assay and a biobank sample, patients received booster mRNA vaccine (initially BNT162b2 per protocol, which was later amended to allow for booster mRNA-1273 vaccine after the Food and Drug Administration (FDA) authorized “booster” doses in fall of 2021). Patients who had received Ad26.CoV2.S vaccine received a booster BNT162b2 vaccine. The patients then returned for follow-up 4 weeks and 4-6 months after their booster dose and their labs were repeated (Supplementary Fig 1).

#### 4^th^ dose study

We have previously reported preliminary findings of a 56% seroconversion rate for patients with cancer who did not have a detectable immune response after 2 doses[17]. For patients who did not seroconvert after 3 doses or had low antibody response (<1000 AU/mL as determined by our in-house assay, Abbott) we hypothesized whether a ‘mix and match’ strategy with 4^th^ dose of COVID-19 vaccine would induce seroconversion/improved boosting of the humoral antibody responses. To study this, we designed a protocol wherein patients who had received 3 prior doses of mRNA vaccines and had undetectable anti-S antibody or had an anti-S antibody level of <1000 AU/mL measured at least 14 days after 3^rd^ dose, would be randomized to an mRNA vs adenoviral 4^th^ vaccine dose. Responses would be then assessed at 4 weeks after the 4^th^ dose through measurement of anti-S antibody results. We also measured complete blood counts, quantitative immunoglobulin levels (IgG, IgA and IgM), lymphocyte subsets, T-cell responses and neutralization activity at baseline and 4 weeks for each of these patients. Following the implementation of this protocol, the Centers for Disease Control (CDC) published a statement that advised that the mRNA vaccines should be preferentially administered over the adenoviral vaccines given concern over rare side effects such as thrombocytopenia and thrombosis syndrome. Following this advisory, we amended our protocol to allow recruitment in a cohort that would receive a 4^th^ dose of the BNT162b2 vaccine to comply with CDC guidelines (Supplementary Fig 1).

#### Anti-SARS-CoV2 Spike (anti-S) antibody assay

The AdviseDx SARS-CoV-2 IgG II assay was used for the assessment of anti-S IgG antibody. AdviseDx is an automated, two-step chemiluminescent immunoassay performed on the Abbott i1000SR instrument. The assay is designed to detect IgG antibodies directed against the receptor binding domain (RBD) of the S1 subunit of the spike protein of SARS-CoV-2. The RBD is a portion of the S1 subunit of the viral spike protein and has high affinity for the angiotensin converting enzyme 2 (ACE2) receptor on the cellular membrane[18, 19] The procedure, in brief, is as follows. Patient serum containing IgG antibodies directed against the RBD is bound to microparticles coated with SARS-CoV-2 antigen. The mixture is then washed of unbound IgG and anti-human IgG, acridinium-labeled, secondary antibody is added and incubated. Following another wash, sodium hydroxide is added and the acridinium undergoes an oxidative reaction, which releases light energy which is detected by the instrument and expressed as relative light units (RLU). There is a direct relationship between the amount of anti-spike IgG antibody and the RLU detected by the system optics. The RLU values are fit to a logistic curve which was used to calibrate the instrument and expresses results as a concentration in AU/mL (arbitrary units/milliliter). This assay recently has shown high sensitivity (100%) and positive percent agreement with other platforms including a surrogate neutralization assay)[20] and also demonstrated high specificity both in the post SARS-CoV-2 infection and post vaccination settings. The cutoff value for this assay is 50 AU/mL with <50 AU/ml values reported as negative and the maximum value is 50000 AU/mL.

### SARS-CoV-2 Interferon Gamma Release Assay

The EUROIMMUN SARS-COV-2 Interferon Gamma Release Assay (Quan-T-Cell SARS-CoV-2) was used for the assessment of patients’ T-cell response to SARS-CoV-2 antigens through analysis of the production of interferon gamma by patient T-cells after exposure to SARS-CoV-2 specific proteins. The assay does not differentiate between vaccine- or infection-induced T-cell responses. The SARS-CoV-2 IGRA assay is performed in two steps as per manufacturer instructions, and a brief protocol follows. First, patient samples from lithium heparin vacutainers are aliquoted into three separate tubes each. These tubes contain either nothing (‘blank’), general T-cell activating proteins (‘mitogen’), or components of the S1 domain of SARS-CoV-2 (‘SARS-CoV-2 activated’). These samples were incubated at 37 degrees for 24 hours before being centrifuged and the plasma separated and frozen at -80 degrees for later analysis. Samples were then batched to be run as a full 96 well plate along with calibrators and controls. Plasma samples were unfrozen and added to an ELISA plate, which was prepared with monoclonal interferon-gamma binding antibodies, along with calibrators and controls. After incubation at room temperature the plate was washed and biotin-labeled anti-interferon gamma antibody was added to bind the patient interferon gamma bound to the plate. The plate was again incubated before being washed of excess antibody and a streptavidin-bound horseradish peroxidase (HRP) enzyme added, which binds strongly to the biotin-labeled antibodies present. This was again incubated and then washed of excess enzyme before a solution of H_2_O_2_ and TMB (3,3’, 5,5;-tetramethylbenzidine, a peroxide-reactive chromogen) is added and allowed to react in the dark for 20 minutes. The reaction is then stopped through the addition of sulfuric acid and the results read at 450 nM with background subtraction at 650 nM. Results for controls and samples were quantified by the calibration curve generated on the same plate, and results were interpreted as long as controls were within the pre-specified range. Blank results for each specimen set were subtracted from each tube in the set and the mIU/mL for both the mitogen and SARS-CoV-2 activated samples were determined with the calibration curve. Samples with mitogen results below 400 mIU/mL were considered ‘invalid’, as the overall T-cell activity for that set was too low, and excluded from analysis. All other sample sets were interpreted as per manufacturer’s instructions based on the SARS-CoV-2 activated sample results: less than 100 mIU/mL were denoted as negative, and greater than or equal to 100 mIU/mL were denoted as positive.

### Neutralization assays

#### Surrogate Virus Neutralization assay for wild type SARS-CoV-2

The SARS-CoV-2 Surrogate Virus Neutralization Test Kit was used to measure antibodies that inhibit the interaction between viral RBD and ACE2 receptor. This test kit uses purified human ACE2 (hACE2) protein-coated enzyme-linked immunosorbent assay (ELISA) plates and horseradish peroxidase (HRP)-conjugated RBD to monitor the presence of circulating antibodies in samples, including peripheral/capillary blood, serum, and plasma, which block the interaction of RBD-HRP with ACE2 with excellent correlation with the gold standard live virus plaque reduction neutralization test.

The kit contains two key components: RBD-HRP and hACE2. The protein-protein interaction between RBD-HRP and hACE2 is disrupted by neutralizing antibodies against SARS-CoV-2 RBD, if present in a sample. After mixing the sample dilutions with the RBD-HRP solution, components are allowed to bind to the RBD. The neutralization antibody complexed to RBD-HRP remains in the supernatant and is removed during washing, The yellow color of the hACE2-coated wells is determined by the RBD HRP binding to the hACE2-coated wells after incubation with 3,3’,5,5’-tetramethylbenzidine (TMB), followed by a stop solution. After the addition of the stop solution, a light-yellow color results from blocking agents interacting with RBDs and inhibiting hACE2 interactions.

#### Microneutralization assay (MNT)

MNT assays were performed in a biosafety level 3 facility at the Icahn School of Medicine at Mount Sinai (ISMMS) as previously described[21]. Briefly, Vero E6-TMPRSS2 cells were seeded in 96-well cell culture plates at 20,000/well in complete Dulbecco’s Modified Eagle Medium (cDMEM). The following day, heat-inactivated serum samples were serially diluted (3-fold) starting at a 1:10 dilution in 1× MEM (10× minimal essential medium (Gibco), 2□mM L-glutamine, 0.1% sodium bicarbonate (Gibco), 10□mM 4-(2-hydroxyethyl)-1-piperazineethanesulfonic acid (HEPES; Gibco), 100□U□ml–1 penicillin, 100□ug/ml–1 streptomycin (Gibco) and 0.2% bovine serum albumin (MP Biomedicals)) supplemented with 10% fetal bovine serum (FBS). The virus diluted at 10,000 tissue culture infectious dose 50% (TCID_50_) per ml of 1× MEM was added to the serum dilutions and incubated for 1□h at room temperature (RT). After removal of cDMEM from Vero E6 cells, 120□μl/well of the virus - serum mix were added to the cells and plates were incubated at 37□°C for 1□h. Mix was removed and 100□μl/well of each corresponding serum dilutions were added in a mirror fashion to the cell plates. Additional 100□μl/well of 1× MEM 1% FBS (Corning) were added to the cells. Plates were incubated for 48□h at 37□°C and fixed with a 10% paraformaldehyde solution (PFA, Polysciences) for 24□h at 4□°C.

For staining, plates were washed with 200□μl of PBS. Cells were permeabilized with 150□μl/well PBS containing 0.1% Triton X-100 for 15□min at RT. Plates were washed 3x with PBST and blocked with 3% milk (American Bio) in PBST for 1□h at RT. Blocking solution was removed and 100□μl/well of the biotinylated mAb 1C7C7 anti-SARS nucleoprotein antibody (generated at the Center for Therapeutic Antibody Development at the ISMMS) were added at 2 μg/ml for 1□h at RT. Plates were then washed 3x with PBST and the secondary antibody goat anti-mouse IgG–HRP (Rockland Immunochemicals) was added at 1:3,000 in blocking solution for 1□h at RT. Plates were washed 3x with PBST, and SIGMAFAST OPD (o-phenylenediamine, Sigma–Aldrich) was added for 10 min at RT. The reaction was stopped with 50□μl/well 3□M hydrochloric acid to the mixture. Optical density (OD_490_) was measured on an automated plater reader (Sinergy 4, BioTek). The inhibitory dilution 50% were calculated as previously described [22].

#### Statistical analysis

The primary endpoint of the third dose study was to assess the rate of booster induced seroconversion amongst patients who remained seronegative at least 28 days following standard set of FDA authorized COVID-19 vaccinations. We hypothesized that booster dosing would convert at least 30% of the enrolled seronegative patients to seropositive as defined by our institutional Clinical Laboratory Improvement Amendments (CLIA) certified SARS-CoV-2 spike IgG assay (as compared to 10% as our null hypothesis). In a pre-specified analysis, at least 26 evaluable seronegative patients were required to have sufficient power to be able to reach this assessment. A McNemar’s test was used to determine the equality of marginal frequencies for paired nominal data with the aid of a homogeneity of stratum effects (HSE) test to check if the effect was the same across all levels of a stratifying variable[23]. A Wilcox test was used to determine if titers of two paired observations have changed over time subsequently using a Kruskal Wallis test to determine if this difference is associated with another variable. For the 4^th^ dose study, a responder was considered any patient who showed seroconversion from negative anti-S antibody to positive anti-S antibody at 4 weeks after 4^th^ dose or increase in titer to >1000 AU/mL at 4 weeks after the 4^th^ dose. An alpha < 0.05 was considered statistically significant. Correlation between continuous variables was assessed using Spearman’s test. All analyses were performed in R (version 3.6.2). This study was approved by The Albert Einstein College of Medicine Institutional Board Review.

## Results

### Duration of immune responsiveness after third dose of covid vaccine in cancer patients

#### Baseline characteristics

We previously reported outcomes for 88 patients enrolled into this study[17]. Here we present our final results for the complete cohort of 106 patients that were enrolled into this study for assessment of the primary endpoint of response at 4 weeks as well as 47 patients who completed 4-6 month follow-up. The baseline characteristics of this cohort are summarized in **Table 1**. The median age was 68 years (63.25-76.5 years)). Fifty five percent (58/106) of patients were female and 45% (48/106) were male. Our cohort was ethnically diverse and included 34% (36/106) Caucasian, 31% (33/106) African-American, 25% (27/106) Hispanic and 8% (9/106) Asian patients. Majority of patients had received mRNA vaccines at baseline. Sixty-eight percent (72/106) received BNT162b2, 26% (28/106) received mRNA-1273 and 6% (6/106) had received Ad26.CoV2.S. Seventy four percent of patients (78/106) received a booster BNT162b2 vaccine and 26% (28/106) patients received booster mRNA-1273 vaccine. The majority of the patients, 62% (66/106), had a hematologic malignancy and 38% (40/106) had a solid tumor diagnosis. Further breakdown of cancer type and cancer status is summarized in **Table 1**. The majority of patients, 75% (80/106), were being actively treated at the time of receiving their booster vaccine.

**Table 1:** Baseline characteristics for 3rd dose cohort.

| Baseline characteristics | n=106 |
| --- | --- |
| Age (median, IQR) | 68 (63.25-76.5) |
| Sex |  |
| Male | 48(45%) |
| Female | 58(55%) |
| Race |  |
| Caucasian | 36(34%) |
| African-American | 33(31%) |
| Hispanic | 27(25%) |
| Asian | 9(8%) |
| Other | 1(1%) |
| **Previous vaccine given** |  |
| BNT162b2 | 72(68%) |
| mRNA-1273 | 28(26%) |
| Ad26.CoV2.S | 6(6%) |
| **Type of booster vaccine** |  |
| BNT162b2 | 78(74%) |
| mRNA-1273 | 28(26%) |
| **Malignancy category** |  |
| Hematologic malignancy | 66(62%) |
| Solid Malignancy | 40(38%) |
| **Lymphoid/Myeloid/Solid** |  |
| Lymphoid | 55(52%) |
| Myeloid | 11(10%) |
| Solid | 40(38%) |
| **Cancer status** |  |
| Active | 69(65%) |
| Progressive | 3(3%) |
| Recurrent | 3(3%) |
| Relapse | 7(7%) |
| Remission | 24(23%) |
| **On treatment at the time of booster** |  |
| Yes | 80(75%) |
| No | 26(25%) |

#### Serology results

Thirty three percent of the patients (35/106) were seronegative after 2 doses. At 4 weeks following the receipt of the booster vaccine, 57% (20/35) of these patients seroconverted and had a detectable antibody response as demonstrated by anti-S antibody testing, meeting the primary endpoint of our study. The median titer at baseline for the entire cohort was 212.1 AU/mL (IQR 50-2873 AU/mL) and the median titer at 4 weeks for the entire cohort was 9997 AU/mL (IQR 880.7-47063 AU/mL) (**Fig 1A**). The median rise in anti-S titer for patients with hematologic malignancies was 2167 AU/mL (IQR 0-10131 AU/mL) versus 31010 AU/mL (IQR 9531-44464 AU/mL) in patients with solid malignancies (p<0.001). Within the hematologic malignancies, patients with lymphoid cancers had a lower rise in median anti-S titers (1169 AU/mL, IQR 0-8661 AU/mL) compared to those with myeloid malignancies; median anti-S titer 9424 AU/mL (IQR 4381-20444 AU/mL) (p<0.001) (**Fig 1B**).

**Figure 1:**
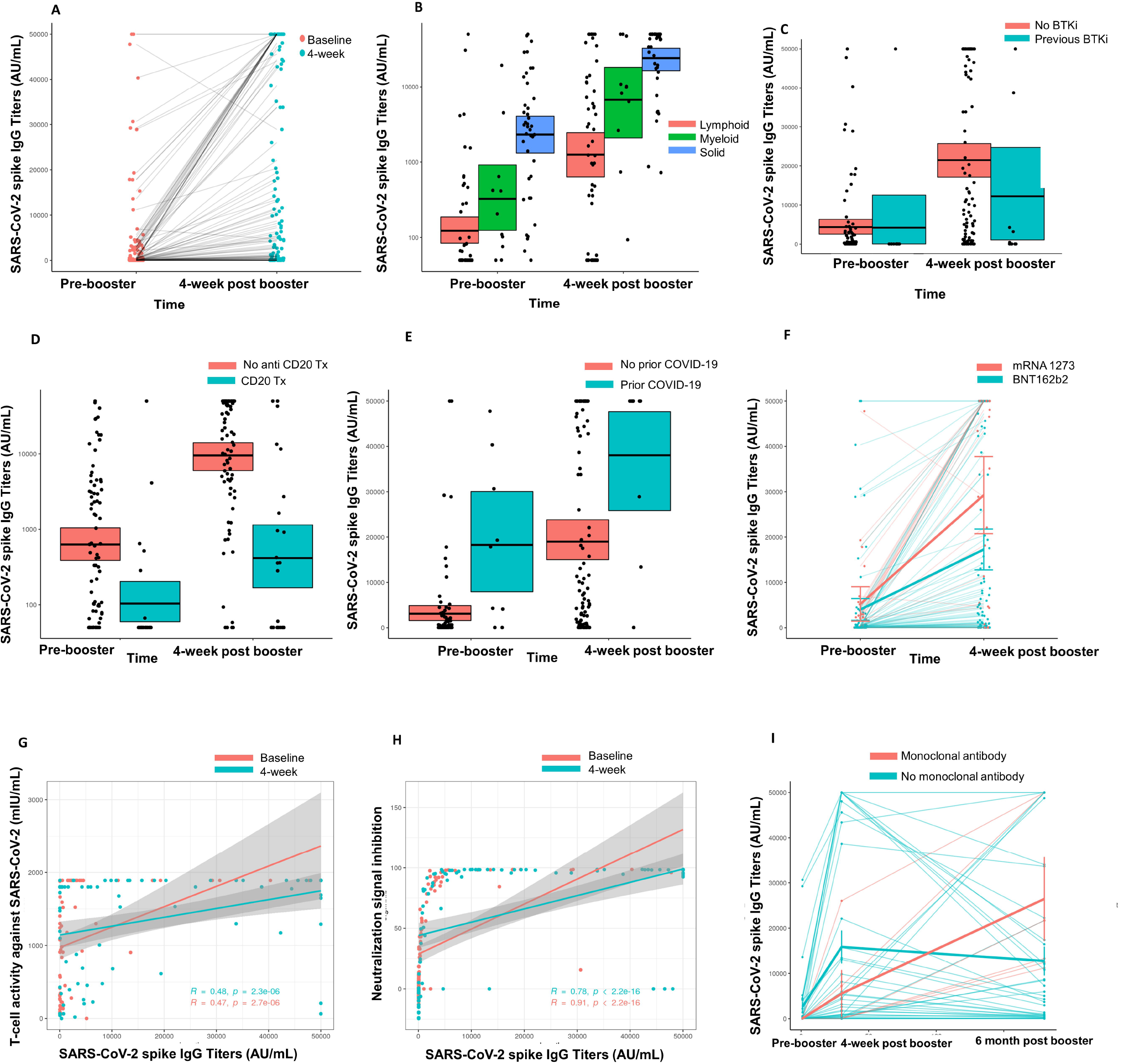
Immunogenicity of 3rd dose of COVID-19 vaccine in seronegative cancer patients. Figure 1A: Figure showing change in anti-S antibody titer at 4 weeks for entire cohort Figure 1B: Figure showing change in anti-S antibody titer at 4 weeks split by cancer type (solid cancer, lymphoid cancer and myeloid cancer) Figure 1C: Figure showing effect of Bruton’s tyrosine kinase inhibitor (BTKi) therapy on anti-S antibody titer at baseline and 4 weeks of 3^rd^ dose Figure 1D: Figure showing effect of anti-CD20 antibody therapy on anti-S antibody titer at baseline and 4 weeks of 3^rd^ dose Figure 1E: Figure showing effect of prior COVID-19 infection on anti-S antibody titer at baseline and 4 weeks of 3^rd^ dose Figure 1F: Figure showing effect of booster type (BNT162b2 vs mRNA 1273) on anti-S antibody titer at baseline and 4 weeks of 3^rd^ dose. Figure 1G: Line diagram showing correlation between anti-Spike IgG titer and baseline T-cell activity at baseline and 4 weeks Figure 1H: Line diagram showing correlation between anti-S titer and signal inhibition for neutralization against WT virus at baseline and 4 weeks. Figure 1I: Anti-Spike IgG titres at baseline, 4 weeks and 6 months after 3^rd^ dose of COVID-19 vaccine in cancer patients. Line shows means with error bars (S.D.).

We further investigated the association of specific anti-cancer therapies with th booster effect. Patients on BTKi therapy (n=12) had a median rise in anti-S antibody of 0 AU/mL (IQR 0-3393 AU/mL) compared to a median rise of 9355 AU/mL (IQR 877.3-34410 AU/mL) in anti-S antibody for patients not on BTKi (p<0.05) (**Fig 1C**). Patients on anti-CD20 antibody therapy (n=25) also had a median rise in anti-S antibody level of 0 AU/mL (IQR 0-910.5 AU/mL) compared to a median rise of 12735 AU/mL (IQR 2842-38863 AU/mL) in patients that did not receive anti-CD20 antibody therapy (p<0.05). (**Fig 1D**). Nine patients had a history of SARS-CoV-2 infection and in this cohort the rise in anti-S titers was higher (median 19350 AU/mL, IQR 9286-32151 AU/mL), compared to those who did not have prior SARS-CoV-2 infection with a median anti-S titer rise, 6706 AU/mL (IQR 444.1-33831 AU/mL). (**Fig 1E**). We also observed that the rise in anti-S titer at 4 weeks was higher for patients who received a mRNA-1273 booster compared to BNT162b2 booster; median 31451 AU/mL vs 5534 AU/mL respectively (**Fig 1F**). This observation was not however, statistically significant. These results are also summarized in **Table 2**.

**Table 2.** Results for 3rd dose of vaccine.

| **Table 2. Results for 3rd dose of vaccine.** |  |  |  |  |
| --- | --- | --- | --- | --- |
| **Spike antibody results** |  | n=106 |  |  |
|  | 4-week negative | 4-week positive | Seroconversion rate | p value |
| Baseline negative | 15 | 20 | 57% | <0.001* |
| Baseline positive | 0 | 71 |  |  |
| Total | 15 | 91 |  |  |
| **Rise in spike antibody titers overall (AU/mL)** | **Median** | **IQR** |  |  |
| Titer at baseline | 212.1 | 50-2873 |  |  |
| Titer at 4 weeks | 9997 | 880.7-47063 |  |  |
| **Rise in spike antibody titers (AU/mL)** | **Median** | **IQR** |  |  |
| Hematologic malignancy | 2167 | 0-10131 |  | <0.001<br>* |
| Solid malignancy | 31010 | 9531-44464 |  |  |
| **Rise in spike antibody titers by solid/lymphoid/myeloid (AU/mL)** |  |  |  |  |
| Lymphoid cancers | 1169 | 0-8661 |  | <0.001<br>* |
| Myeloid cancers | 9424 | 4381-20444 |  |  |
| Solid cancers | 31010 | 9531-44464 |  |  |
| **Association with certain cancer-directed therapies** |  |  |  |  |
| **Bruton's tyrosine kinase inhibitors** |  |  |  |  |
| Change in spike antibody titers (AU/mL) | Median | IQR |  |  |
| Patients on BTKi (n=12) | 0 | 0-3393 |  | <0.001<br>* |
| Patients not on BTKi | 9355 | 877.3-34410 |  |  |
| **Anti-CD20 antibody treatment** |  |  |  |  |
| Change in spike antibody titers (AU/mL) | Median | IQR |  |  |
| Patients on CD20 (n=25) | 0 | 0-910.5 |  | 0.0133* |
| Patients not on CD20 | 12735 | 2842-38863 |  |  |
| **Anti-CD20 antibody treatment within 6 months** | Median | IQR |  |  |
| Yes | 0 | 0-0 |  | 0.0548<br>2 |
| No | 587 | 0-4314 |  |  |
| **Change in spike antibody titer by prior COVID infection** | Median | IQR |  |  |
| Yes (n=9) | 19350 | 9286-32151 |  | 0.3051 |
| No (n=96) | 6706 | 444.1-33831 |  |  |
| **Change in spike antibody titer by type of booster given** | Median | IQR |  |  |
| BNT162b2 | 5534 | 433.8-18074 |  | 0.0901<br>4 |
| mRNA-1273 | 31451 | 515.5-45057 |  |  |
| **T-cell activity** |  |  |  |  |
| Baseline | n=88 | % |  |  |
| Positive | 65 | 74% |  |  |
| Negative | 23 | 26% |  |  |
| **4-week** | n=89 |  |  |  |
| Positive | 76 | 85% |  |  |
| Negative | 13 | 15% |  |  |
| **Baseline Neutralization activity assay (all evaluable patients, WT virus)** |  |  |  |  |
|  | Anti-S antibody negative | Anti-S antibody positive | Total | p value |
| Neutralizing antibodies detected | 0 | 47 | 47 | <0.001 |
| Neutralizing antibodies not detected | 35 | 21 | 56 |  |
| Total | 35 | 68 | 103 |  |
| **Four-week Neutralization activity assay (all evaluable patients, WT virus)** |  |  |  |  |
|  | Anti-S antibody negative | Anti-S antibody positive | Total | p value |
| Neutralizing antibodies detected | 0 | 77 | 77 | <0.001 |
| Neutralizing antibodies not detected | 15 | 8 | 23 |  |
| Total | 15 | 85 | 100 |  |
| **Four week Neutralization assay (seronegative cohort 4 weeks)** | n=35 |  |  |  |
| **Wild type** |  |  |  |  |
| Negative | 19 | 54% |  |  |
| Positive | 16 | 46% |  |  |
| **Omicron** |  |  |  |  |
| Negative | 29 | 83% |  |  |
| Positive | 6 | 17% |  |  |

#### T cell immune responses

We also studied T-cell immune responses through a SARS-CoV-2 interferon gamma release assay. At baseline, 88 patients had evaluable T-cell results and a positive T cell response against SARS-CoV-2 was seen in 74% (65/88) patients. Of these, 21 patients were seronegative for anti-S antibody at baseline. At 4 weeks, 89 patients had evaluable T-cell results and a positive result was seen in 85% (76/89) percent patients. Of the 15 patients with negative anti-S antibody at 4 weeks, 11 had a positive T-cell response. Fourteen patients who had a negative T-cell assay response at baseline had a positive T-cell response at 4 weeks. Anti-S titer showed a positive correlation with T-cell response at baseline and at 4 weeks for this cohort (p<0.001) (**Fig 1G**). These results are summarized in **Table 2**.

#### Neutralization assays

##### Neutralization assay against wild type virus

We tested neutralization pre- and post-3^rd^ dose in this cohort using the GenScript surrogate virus neutralization assay. At baseline, biobanked samples from 103 patients were tested for neutralizing antibodies. Of these, 35 patients were seronegative at baseline and 68 patients were seropositive. Neutralizing antibodies were detected in 47 of 68 (69%) patients who were seropositive at baseline. The correlation between seropositivity and presence of neutralizing antibodies was statistically significant (p<0.001, Fisher’s exact test).

At 4 weeks post 3^rd^ dose, samples from 100 patients were available for testing. Eighty five of these patients were seropositive at 4 weeks and 15 were seronegative. Neutralizing antibodies were detected in 77 of 85 (91%) seropositive patients at 4 weeks. The correlation between seropositivity and presence of neutralizing antibodies was also statistically significant at 4 weeks (p<0.001, Fisher’s exact test).

We also analyzed the correlation of anti-S titers at baseline and 4 weeks to the percentage of virus neutralization, with 30% or more neutralization being consistent with positive result for detection of neutralizing antibodies. We observed that at baseline and 4 weeks, anti-S titers correlated with percentage of viral neutralization with higher titers correlating with higher percentage of viral neutralization (Figure **1H** <0.001 by Spearman rank correlation). These results are summarized in **Table 2**.

##### Neutralization against Omicron BA.1

Thirty-five patients were found be seronegative after the 3^rd^ dose. Due to the emergence of the Omicron BA.1 wave, we further assessed neutralization activity for the seronegative cohort (N=35) against wild type (WT) SARS-CoV-2 and BA 1.1.529 (Omicron BA.1). At 4 weeks neutralization was noted in 46% patients (16/35) for the WT virus while only 17% of patients had detectable neutralization activity (6/35) for the Omicron variant. These results are summarized in **Table 2**.

#### Six-month follow-up post 3^rd^ dose of vaccine

Forty seven patients (44%) out of 106 completed 4-6 months follow-up for the third dose study. All these patients were seropositive 4 weeks after the 3^rd^ dose and strikingly, we observed that all patients maintained a positive anti-S antibody at 4-6 months follow-up. Eleven of these 47 patients had solid malignancies and 36 had hematologic malignancies. Six patients had received anti-Covid monoclonal antibody (moAb) therapy as per standard of care (4 tixagevimab-cilgavimab or Evusheld, 1 casirivimab/imdevimab or regen-co-v, and 1 sotrovimab between the 4-week and 4-6 months follow-up. A striking increase in titers in this small cohort of patients was noted to a median titer of 17481.2 AU/mL. Four patients had breakthrough SARS-CoV-2 infections and 9 patients had received a 4^th^ dose of COVID-19 vaccine outside of the context of the study prior to the time of 4-6 month follow-up. The median decline in titer for 41 patients who did not receive anti-SARS-CoV-2 moAb treatment in the interim to confound results was -922.2 AU/mL. When compared to the antibody levels 4 weeks after booster vaccination, the median percentage decline in titers was 56.4%. However, despite the noted decline not a single patient in this cohort seroreverted **(Fig 1I)**, especially when compared to decline post 2 vaccines. In our initial report of seroconversion post-3^rd^ booster, we reported waning of immunity in 99 patients post 2 vaccines. The median decline in the 99 patient cohort was 72.1% with 2 patients losing detectable antibody response [17]

### Efficacy of 4^th^ dose vaccine for patients that were seronegative or low seropositive after 3^rd^ dose

#### Baseline characteristics

Eighteen patients were enrolled into the 4^th^ dose study. Median age for this cohort was 69.5 years (IQR 65.5-73.8). Thirty nine percent (7/18) were seronegative at baseline and 61% patients (11/18) were sero-low (anti-S ab <1000 AU/mL). All patients had hematologic malignancies in this cohort and the breakdown of diagnoses is provided in **Table 3**. Eighty three percent of the patients (15/18) received BNT162b2 4^th^ booster shots and 17% (3/18) patients received Ad26.CoV2.S as their 4^th^ booster vaccine. In addition, we also measured baseline complete blood counts (CBC), lymphocyte subsets, immunoglobulin G, A and M (quantitative Ig) levels at baseline and 4 weeks. (**Table 4**).

**Table 3:**
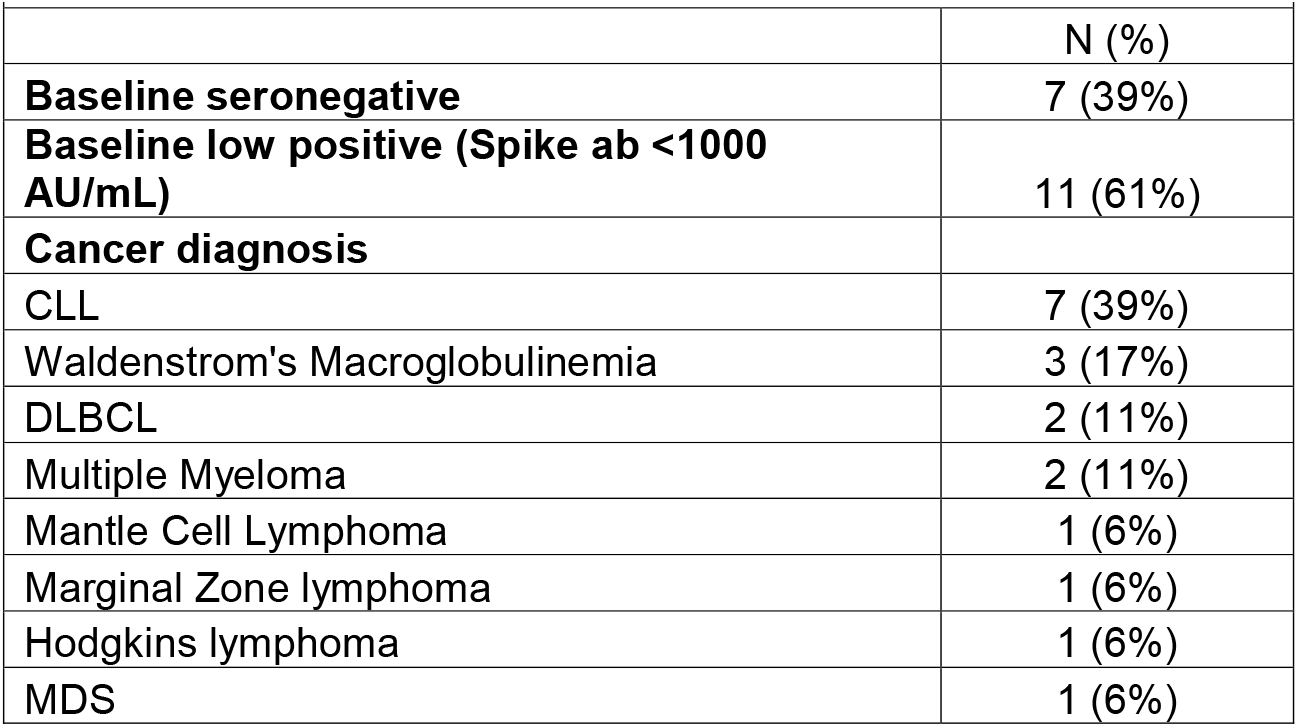
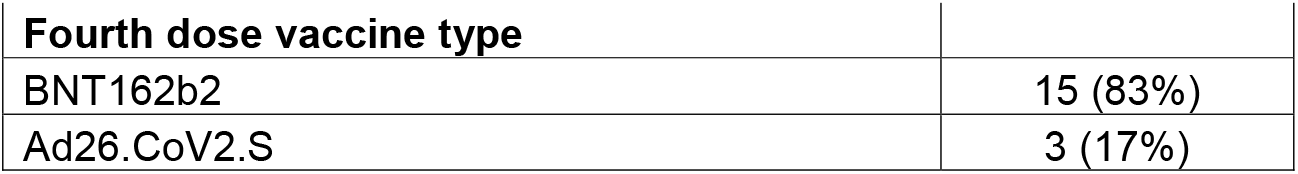
Baseline characteristics of the 4th dose cohort.

| **Table 3: Baseline characteristics of the 4th dose cohort.** |  |
| --- | --- |
|  | N (%) |
| **Baseline seronegative** | 7 (39%) |
| **Baseline low positive (Spike ab <1000 AU/mL)** | 11 (61%) |
| **Cancer diagnosis** |  |
| CLL | 7 (39%) |
| Waldenstrom's Macroglobulinemia | 3 (17%) |
| DLBCL | 2 (11%) |
| Multiple Myeloma | 2 (11%) |
| Mantle Cell Lymphoma | 1 (6%) |
| Marginal Zone lymphoma | 1 (6%) |
| Hodgkins lymphoma | 1 (6%) |
| MDS | 1 (6%) |

| Fourth dose vaccine type |  |
| --- | --- |
| BNT162b2 | 15 (83%) |
| Ad26.CoV2.S | 3 (17%) |

**Table 4:** Correlation of 4th dose vaccine response with baseline characteristics.

|  |  |  |  |
| --- | --- | --- | --- |
|  | **Non-responder (n=6)** | **Responder (n=12)** | **p value** |
| Age | 79.5 | 67.5 | 0.01293 * |
| Baseline WBC | 4.95 | 5.15 | 0.45 |
| Baseline ANC | 2.6 | 3.5 | 0.26 |
| Baseline ALC | 1.2 | 1.3 | 0.57 |
| Baseline AMC | 0.5 | 0.65 | 0.73 |
| Baseline Absolute CD3 | 773 | 835.5 | 0.57 |
| Baseline Absolute CD4 | 406.5 | 407.5 | 0.71 |
| Baseline Absolute CD8 | 310 | 247 | 0.40 |
| Baseline Absolute CD19 | 1 | 113.5 | 0.04874 * |
| Baseline Absolute CD16/56 | 243.5 | 200 | 0.57 |
| Baseline IgG | 777 | 757 | 0.51 |
| Baseline IgA | 90.5 | 118 | 0.57 |
| Baseline IgM | 17 | 60.5 | 0.001442 * * |
| 4 week WBC | 5.1 | 5.8 | 0.40 |
| 4 week ANC | 2.7 | 3.45 | 0.57 |
| 4 week ALC | 1.1 | 1.4 | 0.60 |
| 4 week AMC | 0.55 | 0.65 | 0.60 |
| 4 week absolute CD3 | 754 | 983 | 0.40 |
| 4 week absolute CD4 | 461.5 | 369.5 | 0.93 |
| 4 week absolute CD8 | 297.5 | 269 | 0.40 |
| 4 week absolute CD19 | 2.5 | 105 | 0.07 |
| 4 week absolute CD16/56 | 232.5 | 219 | 0.93 |
| 4 week IgG* | 741.5 | 832 | 0.62 |
| 4 week IgA* | 86 | 112 | 0.69 |
| 4 week IgM* | 15 | 62 | 0.003561 * * |
| *n=11 for responder group |  |  |  |

#### Anti-spike IgG responses after the 4^th^ dose

A patient was classified as a responder if they 1) had positive anti-S antibody at 4 weeks if seronegative at baseline or 2) if they achieved a titer of >1000 AU/mL at 4 weeks if they were sero-low at baseline. As such, we observed a 67% response rate (12/18) in patients for the 4^th^ dose cohort. Two of 7 seronegative patients seroconverted to positive anti-S antibody at 4 weeks with a seroconversion rate of 29% in this cohort. All low seronegative patients (11/11) responded with an IgG level > 1000 after the 4^th^ dose (**Fig 2A**). For the whole cohort, the median anti-S antibody at baseline was 131.1 AU/mL (<50-432.9 AU/mL) and at 4 weeks was 1700 AU/mL (IQR 64.3-18627 AU/mL). We further investigated association of baseline laboratory values, such as CBC, lymphocyte subsets and quantitative Ig levels and observed that patients in the responder group had higher baseline IgM (60.5 mg/dl) compared with the non-responder group (median 17 mg/dl, p<0.001) (**Fig 2B**). Additionally, we also observed that the median CD19+ cell count was significantly lower in the non-responder group versus the responder group (1 vs 113, p=0.04). These results are summarized in **Table 5**.

**Figure 2:**
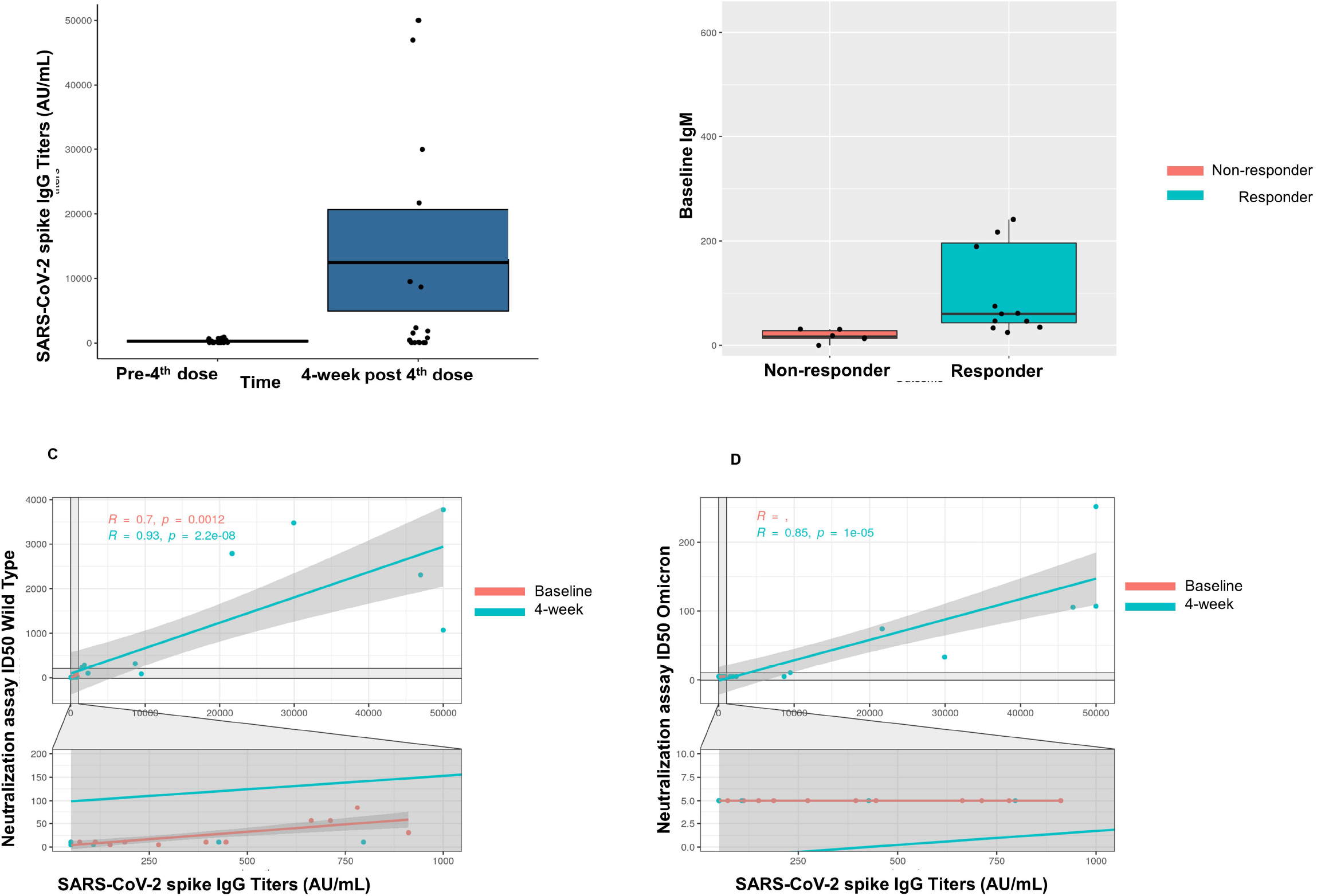
Immunogenicity of the 4^th^ dose of COVID-19 vaccine in cancer patients with seronegativity after 3 doses. Figure 2A: Anti-spike IgG levels after the 4^th^ dose of COVID-19 vaccine for the entire cohort Figure 2B: Correlation of baseline IgM levels with response to 4^th^ dose of vaccine Figure 2C: Line diagram showing correlation between anti-S titer and neutralization activity for WT virus at baseline and 4 weeks Figure 2D: Line diagram showing correlation between titer and neutralization activity for Omicron strain at baseline and 4 weeks

**Table 5:** Results for 4th dose study.

| **Table 5: Results for 4th dose study.** |  |  |  |
| --- | --- | --- | --- |
| **Overall response** | 18 |  |  |
| Responder | 12 | 67% |  |
| Non-responder | 6 | 33% |  |
| **Median age** |  | IQR |  |
| Responder | 67.5 | 63.75- 70.75 | 0.01293 * |
| Non-responder | 79.5 | 72.75- 81.75 |  |
| **Median baseline IgM** |  |  |  |
| Responder | 60.5 |  | 0.001442<br>* * |
| Non-responder | 17 |  |  |
| **Median spike antibody at baseline (AU/mL)** | 131.1 | <50-432.9 |  |
| **Median spike antibody at 4 weeks (AU/mL)** | 1700 | 64.3-18627 |  |
| **T-cell activity at baseline** | n=14 |  |  |
| Positive | 11 | 79% |  |
| Negative | 3 | 21% |  |
| **T cell activity at 4 weeks** | n=18 |  |  |
| Positive | 17 | 94% |  |
| Negative | 1 | 6% |  |
| **Baseline** |  |  |  |
| **Neutralization assay baseline** | Negative | Positive |  |
| WT | 6(33%) | 12(67%) |  |
| Omicron | 18(100%) | 0(0%) |  |
| **Neutralization assay 4 week** | Negative | Positive |  |
| WT | 5(28%) | 13(72%) |  |
| Omicron | 12(67%) | 6(33%) |  |

#### T Cell activity against SARS-CoV-2 after the 4^th^ dose

T-cell activity was assessed at baseline and at 4 weeks using the SARS-CoV-2 interferon gamma release assay. At baseline, 14 patients had evaluable T-cell responses and a positive response was noted in 79% patients (11/14). Of these, 3 patients had negative anti-S antibody at baseline. At 4 weeks after the 4^th^ dose, a positive T-cell response was seen in 17/18 (94%) patients. These results are summarized in **Table 5**.

#### Neutralization activity against SARS-CoV-2 after 4^th^ dose

We also assessed neutralization activity at baseline and at 4 weeks against WT and Omicron (B 1.1.529, BA.1). The results are summarized in **Table 5**. Overall, neutralization activity was seen in 67% patient samples at baseline and in 72% patient samples at 4 weeks. Strikingly, neutralization activity against Omicron was absent in all patient samples at baseline, however became detectable in 33% (6/18) patients at 4 weeks after the 4^th^ dose. The titer of anti-S antibody correlated with neutralization activity at baseline and at 4 weeks against the WT virus (p<0.001) (**Fig 2C**). We also observed correlation between the titer of anti-S antibody with neutralization activity at 4 weeks for the Omicron variant (**Fig 2D**).

#### Exploratory analysis for immunoglobulin levels

The observation for baseline IgM correlating with response to the 4^th^ dose of the COVID-19 vaccine led us to perform an exploratory analysis to assess if IgG and IgA levels would also correlate with the response. Given that our 4^th^ dose cohort was small, we performed this exploratory analysis by combining the baseline immunoglobulin levels for the baseline seronegative cohort for the 3^rd^ dose study (n=35) and baseline immunoglobulin levels for the 4^th^ dose study (n=18). In this exploratory analysis, we observed that the median levels for all immunoglobulin subtypes were lower in patients who either did not seroconvert after the 3^rd^ dose or did not respond to the 4^th^ dose (IgA 49 mg/dL vs 116.5 mg/dL, IgM 16.6 mg/dL vs 48.3 mg/dL, IgG 488 mg/dL vs 759.5 mg/dL with p values of 0.05, 0.002 and 0.006 respectively (Kruskal Wallis test).

## Discussion

Since the authorization of 3^rd^ doses for patients with a weakened immune system, several studies have shown enhanced immunogenicity for a third dose of COVID-19 vaccine in patients with cancer. [17, 24, 25] In particular, patients with lymphoid malignancies have been consistently shown to have reduced seroconversion after 2 doses of the COVID-19 vaccines[26-29]. Studies looking at immunogenicity of a 3^rd^ dose of COVID-19 vaccines have reported that a subset of these patients can be induced to have an immune response with the 3^rd^ dose of the COVID-19 vaccines.[17, 30]

Correlation between anti-S antibody titers and neutralization activity in patients with cancer has been demonstrated [31]. However, the emergence of the Omicron (B.1.1.529) variant which was discovered in November 2021 and then spread quickly globally, the situation changed. Omicron, with its extensive mutations in neutralizing epitopes, is able to at least partially evade *in vitro* neutralizing antibodies induced by 3^rd^ doses in patients with cancer[31, 32]. The potential utility and timing of a 4^th^ COVID-19 vaccine dose has been brought up especially for those who are at risk for poor seroconversion after 3^rd^ doses[33], with the Centers for Disease Control (CDC) recommending 2 additional boosters following a 3-vaccine primary series[34]. These variants in part overcome vaccine-induced immunity and are resistant to many of the available monoclonal antibody products[31, 32, 35].

Our results demonstrate that a 3^rd^ dose of COVID-19 vaccine boosts detectable anti-SARS-CoV-2 immunity in the majority of cancer patients and can seroconvert a subset of them not responding to prior vaccine doses. The 3^rd^ COVID-19 vaccine also results in boosting of T-cell responses and leads to a rise in neutralizing antibodies. Patients who have received anti-CD20 antibody therapy or BTK inhibitors remain at risk for lower seroconversion whereas those who have been infected with COVID-19 in the past, have a very strong immune response likely due to immunologic memory. Our results show that the higher the titer of the anti-S antibody, the higher likelihood of neutralization in a surrogate neutralization assay adding to the evidence that this may be a good strategy to prevent symptomatic SARS-CoV-2 infection as well as an appropriate surrogate marker to guide research and clinical management [36]. Our study also provides the reassuring finding that the large majority of patients with cancer retain detectable humoral immunity at 6 months post 3^rd^ dose of COVID-19 vaccination.

Reports of efficacy of 4^th^ COVID-19 vaccine doses are emerging. A study from Israel demonstrated enhanced Omicron neutralization after a 4^th^ dose of COVID-19 mRNA vaccine in healthy health-care workers[37]. However a study of 25 patients with solid organ transplant recipients showed that the 4^th^ dose was not effective in inducing Omicron neutralization[38]. Such a study has not been published yet for patients with cancer, making this an unmet need. We designed a prospective cohort study of a 4^th^ dose of the COVID-19 vaccine in patients with cancer precisely to address this question. Our results suggest that in cohorts of highly immune suppressed patients, baseline assessment of immunity based on prior treatment history and immunological markers such as IgM levels and CD19+ cell levels may help predict the response to COVID-19 booster vaccinations and support administration of additional vaccine doses even in highly immune suppressed individuals. Notably, serum IgM levels were previously shown to correlate with mRNA vaccine responses of solid organ transplant recipients[39]. In addition, further testing to assess serological and cellular markers of the response may be helpful to identify the patients at highest risk to prioritize these patients for preventive/prophylactic strategies as well as enrichment markers for further experimental studies. Finally, the 4^th^ vaccine dose results in a significant increase in anti-spike antibodies in low seropositive patients and seroconversion in a proportion of seronegative immunosuppressed patients with cancer. Similar to previous reports, the booster doses do lead to enhanced neutralization activity against the WT virus, but not the Omicron (BA.1) variant. Future efforts are needed to evaluate variant-specific vaccines as well as additional protective measures, such as passive immunization strategies, especially for this immunosuppressed patient population that may not benefit as much as healthy controls from booster doses of existing vaccines.

## Data Availability

All data produced in the present study are available upon reasonable request to the corresponding author

## Acknowledgements

This study was supported with funding from the National Cancer Institute Grant 3P30CA013330-49S3 and NCORP Grant 2UG1CA189859-06. The authors also acknowledge support from the Jane and Myles Dempsey Family. The funders had no role in study design, data collection and analysis, decision to publish, or preparation of the manuscript. Work in the Krammer laboratory was partially funded by the Centers of Excellence for Influenza Research and Surveillance (CEIRS, contract # HHSN272201400008C), the Centers of Excellence for Influenza Research and Response (CEIRR, contract # 75N93021C00014), by the Collaborative Influenza Vaccine Innovation Centers (CIVICs contract # 75N93019C00051) and by institutional funds. Finally, this effort was also supported by the Serological Sciences Network (SeroNet) in part with Federal funds from the National Cancer Institute, National Institutes of Health, under Contract No. 75N91019D00024, Task Order No. 75N91020F00003. The content of this publication does not necessarily reflect the views or policies of the Department of Health and Human Services, nor does mention of trade names, commercial products or organizations imply endorsement by the U.S. Government.

## Author contributions

Conception and design: AT, LCS, LAP, BH, AV. Patient referral recruitment and follow-up: AT, LCS, BD, IM. Data curation: AT, LCS. Laboratory investigations: JMCQ, SS, VT, SC, TDB, JR, RO, MS. Logistical support : ML, RQ. Administrative support: EC, BW, FK, Data analysis: KP, All authors contributed to the writing of the manuscript.

