## Supplemental Figure for "Efficacy and longevity of immune response to 3^rd^ COVID-19 vaccine and effectiveness of a 4^th^ dose in severely immunocompromised patients with cancer"

### Slide 1
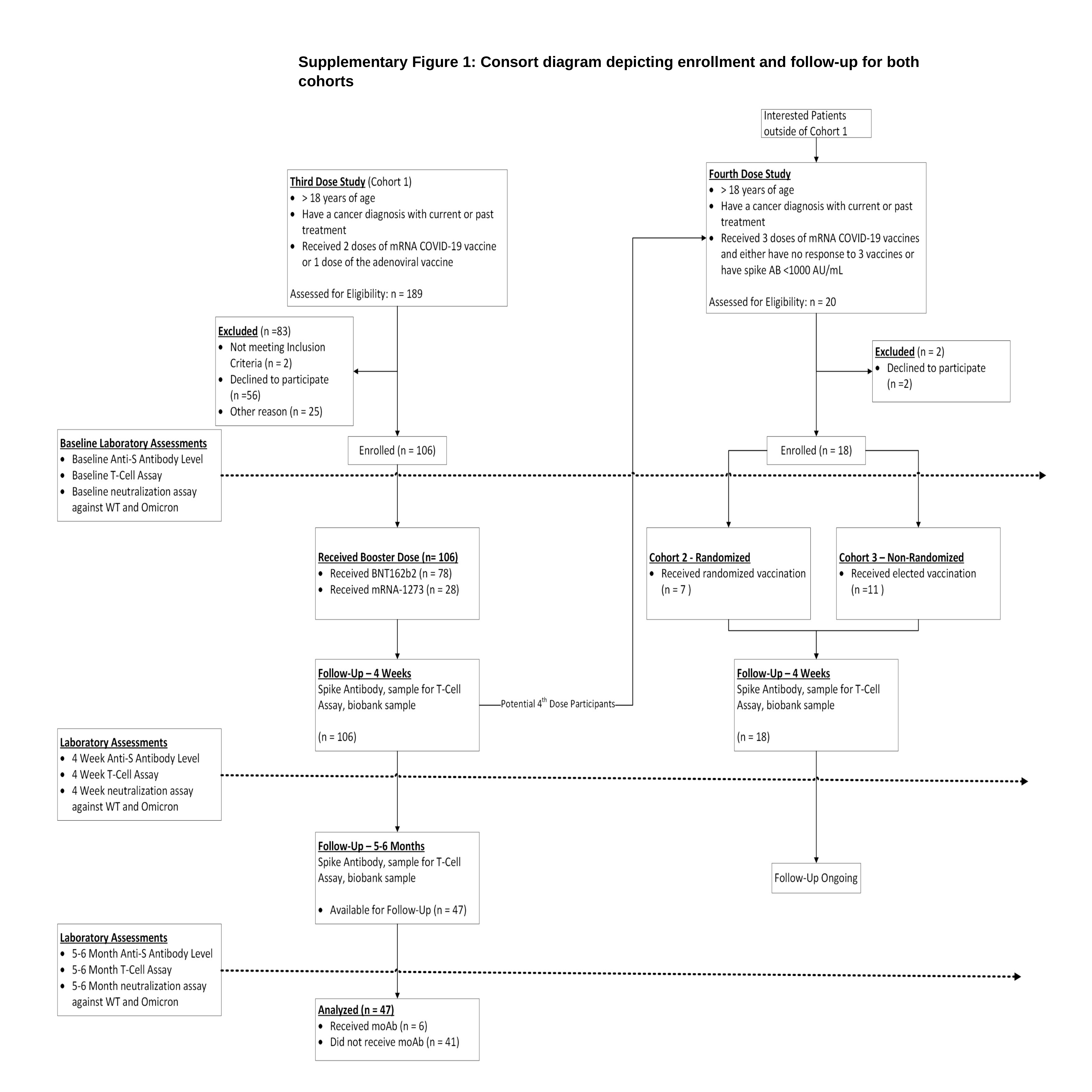

Figure 1
Supplementary Figure 1: Consort diagram depicting enrollment and follow-up for both cohorts
